# A Novel Machine Learning Based Framework for Developing Composite Digital Biomarkers of Disease Progression

**DOI:** 10.1101/2024.09.23.24313737

**Authors:** Song Zhai, Andy Liaw, Judong Shen, Yuting Xu, Vladimir Svetnik, James J. FitzGerald, Chrystalina A. Antoniades, Dan Holder, Marissa F. Dockendorf, Jie Ren, Richard Baumgartner

## Abstract

**Background:** Current methods of measuring disease progression of neurodegenerative disorders, including Parkinson’s disease (PD), largely rely on composite clinical rating scales, which are prone to subjective biases and lack the sensitivity to detect progression signals in a timely manner. Digital health technology (DHT)-derived measures offer potential solutions to provide objective, precise, and sensitive measures that address these limitations. However, the complexity of DHT datasets and the potential to derive numerous digital features that were not previously possible to measure pose challenges, including in selection of the most important digital features and construction of composite digital biomarkers.

**Methods:** We present a comprehensive machine learning based framework to construct composite digital biomarkers for progression tracking. This framework consists of a marginal (univariate) digital feature screening, a univariate association test, digital feature selection, and subsequent construction of composite (multivariate) digital disease progression biomarkers using Penalized Generalized Estimating Equations (PGEE). As an illustrative example, we applied this framework to data collected from a PD longitudinal observational study. The data consisted of Opal^TM^ sensor-based movement measurements and MDS-UPDRS Part III scores collected at 3-month intervals for 2 years in 30 PD and 10 healthy control participants.

**Results:** In our illustrative example, 77 out of 235 digital features from the study passed univariate feature screening, with 11 features selected by PGEE to include in construction of the composite digital measure. Compared to MDS-UPDRS Part III, the composite digital measure exhibited a smoother and more significant increasing trend over time in PD groups with less variability, indicating improved ability for tracking disease progression. This digital composite measure also demonstrated the ability to classify between de novo PD and healthy control groups.

**Conclusion:** Measures from DHTs show promise in tracking neurodegenerative disease progression with increased sensitivity and reduced variability as compared to traditional clinical scores. Herein, we present a novel framework and methodology to construct composite digital measure of disease progression from high-dimensional DHT datasets, which may have utility in accelerating the development and application of composite digital biomarkers in drug development.

## 1 Introduction

Neurodegenerative diseases, including Parkinson’s Disease (PD), are an area of vast unmet medical need. Drug development efforts in this area have increasingly focused on the search for disease-modifying therapies that slow down the underlying disease progression mechanisms. However, a lack of validated measures that allow for disease progression to be monitored objectively, relatively rapidly, and with high precision makes it challenging to effectively demonstrate therapeutic efficacy and hinders drug development efforts. PD clinical trials generally use the Movement Disorder Society - Unified Parkinson’s Disease Rating Scale (MDS-UPDRS) to track disease progression longitudinally. However, MDS-UPDRS is subjective in nature, relies on patient and caregiver-reported symptoms and clinician’s qualitative ratings [1], is slow to change, and has low measurement precision, resulting in large and lengthy clinical trials to test efficacy for potential disease modifying therapies [2].

Recent advances in digital health technologies (DHTs) offer unprecedented opportunities to collect more objective, precise, and sensitive measures, both in the clinic and remotely, that were out of reach in the past. Such measures could provide new insights into neurogenerative disease progression, including for Parkinson’s disease. There are many studies that have investigated using measures from sensor-based digital health technologies in neurodegenerative diseases [3 – 11]. These studies have collectively demonstrated that many neurodegenerative disease symptoms can be quantified by DHTs. Moreover, multiple longitudinal observational studies have shown that digital measures can pick up changes over time that are indicative of disease progression [12 – 18]. It is further thought that the objective measures enabled by DHTs could offer improved sensitivity and reduced variability [12, 19], which could translate to smaller and shorter clinical trial designs [20] and, in turn, potential for accelerated drug development. Despite promising results, the longitudinal studies published to date have used different DHTs and analysis methodologies to identify the digital features of importance and to derive respective digital clinical measures, making it difficult to compare across studies or create consensus among the research community. Open discussions on the methodology of digital clinical measure development and evaluation are critically needed to move the field forward.

It has been increasingly recognized that composite digital measures, rather than reliance on individual digital features, are needed for more effective measurement of disease progression as compared to traditional clinical composite scores. Adams et al. [21] showed that no individual digital feature (from gait, tremor, turns, speech, and cognition) outperformed MDS-UPDRS Part III (a composite clinical score) in terms of the standardized change from baseline after 12 months in a PD observational study (WATCH-PD). Furthermore, Czeck et. al. [28] demonstrated individual sensor-based digital features of upper and lower extremity bradykinesia often lacked strong sensitivity to longitudinal changes, whereas digital composite scores showed significant differences over 12 months in WATCH-PD.

There have been several examples where composite digital measures were developed for disease classification and/or tracking symptom progression [22 - 30]; however, the approach taken has varied, and there have been limited discussions on the methodologies to effectively select informative digital features and construct the most performant composite measures. For example, Perumal and Sankar [22] developed a Linear Discriminant Analysis (LDA) classifier using multiple gait features collected from wearable sensors to distinguish between PD patients and healthy control (HC) subjects. Czech et al. [28] constructed composite digital scores using pre-defined combinations of features from single tasks (pronation-supination and toe-tapping) and used them to measure longitudinal progression of bradykinesia after 1 year. Sotirakis et al. [30] developed a Random Forest model to estimate the MDS-UPDRS III values using gait and sway features and used the model to detect progression of motor symptoms longitudinally. These efforts vary in terms of the measure construction (pre-defined vs. supervised ML, choice of models), the clinical label selection (MDS-UPDRS III total score or single item), the selection of digital tasks (single task e.g., toe-tapping or a combination of tasks), as well as the selection of input features (e.g., whether features are pre-screened). Overall, the field has not adopted consistent and systematic methods and/or analysis frameworks. Therefore, there is an urgent need to develop methodologies and analysis pipelines for the construction of composite digital measures for disease progression tracking, tailored for high-dimensional, longitudinal data with digital features collected from sensor technologies.

The types of data generated by DHTs are often longitudinal and high dimensional, which differs from traditional clinical measures, calling for novel analytical strategies to handle such data for the construction of composite digital measures. Unlike traditional clinical measures that collect a defined set of measures at each time point, DHTs leverage various sensors to generate large amounts of time-series data (e.g., acceleration, screen touch, audio/video, keyboard press), either collected from defined active task-based assessments or from passive monitoring. Such data are often not readily analysable statistically and need to be aggregated and transformed into digital features first. For example, for measurement of physical activity, continuous accelerometer signals are often converted to epoch level activity counts and then aggregated over time into features such as daily total activity count, total steps, non-sedentary time, etc., for further statistical analysis. There can be large numbers of features derived from the high-frequency sensor signals; such features may have various data types (i.e., categorical, continuous, duration, etc.) and clinimetric properties, many of which may not yet have been fully explored as it was not previously possible to measure them without use of DHTs. These features could have intrinsic skewness in distribution, floor/ceiling effects, as well as unknown redundancies and covariances. In addition, the high frequency nature of DHT data collection and potential for remote data acquisition can also lend itself to higher levels of data missingness. Furthermore, not all digital features that can be generated from sensor data may have clinical significance or be valuable for creating composite digital measures. These attributes of DHT data make it a unique challenge in the development of composite digital measures to track longitudinal disease progression.

Machine learning (ML) methods offer a valuable tool for selecting the most informative digital features to reflect disease progression and to construct clinically meaningful composite digital measures. ML-based techniques can often improve prediction performance in analysing digital data in neurodegenerative diseases; however, existing ML methodologies for longitudinal data analysis are also challenged by the high dimensionality of DHT data. For example, although the generalized estimating equations (GEE) method [31] incorporating different patterns of working correlation matrix across multiple timepoints has been widely used in longitudinal data analysis, the direct use of classical unpenalized GEE in high-dimensional longitudinal data analysis may lead to misleading results [32]. To address this, an ML-based penalized GEE (PGEE) method [32] could be used to improve upon the GEE method in handling DHT data. PGEE performs simultaneous coefficient estimation and variable selection for longitudinal data analysis with high-dimensional covariates by including a penalty term in the GEE model, which can be better-suited to handle high-dimensional feature sets.

In this paper, we propose a principled and comprehensive methodology framework for the development of novel composite digital biomarkers, derived from DHT data and anchored to the MDS-UPDRS score, to measure neurodegenerative disease progression. This framework includes data processing, univariate digital feature screening, multivariate (composite) digital biomarker construction (using PGEE methods), and composite biomarker performance evaluation.

We further demonstrate the utility of this framework by applying it to a sample dataset containing high-dimensional, longitudinal movement data collected by a body-worn accelerometer system from a PD longitudinal observation study. The current analytical challenges of high-dimensional and longitudinal digital data and path forward for the application of composite digital biomarkers in measurement of neurodegenerative disease progression are also discussed.

## 2 Materials and Methods

### 2.1 Study Overview

To illustrate our proposed methodology to construct composite digital measures for tracking longitudinal disease progression, we applied the framework to data from 30 PD patients (10 de novo PD patients, 10 mild-to-moderate PD patients on levodopa, and 10 advanced PD patients) and 10 healthy control subjects from a PD longitudinal observational study conducted at John Radcliffe Hospital in Oxford, UK [11, 29, 33]. The participants visited the clinic once every 3 months for 2 years. At each visit, they wore six synchronized inertial measurement units (IMUs) (“Opal” sensors, APDM Wearable Technologies, a Clario Company) across their body and performed two-minute walk, postural sway, and timed up-and-go (TUG) tasks. The Mobility Lab^TM^ software (APDM Wearable Technologies, a Clario Company) was then used to process these raw sensor signals, and generate epoch-level digital features at each instance of a time period or physical movement (e.g., per minute, per step, per turn, or per sit-to-stand event). The MDS-UPDRS Part III assessments were also conducted at these clinic visits. The MDS-UPDRS Part III score and subscales (including Bradykinesia, Postural and Gait, Rigidity, and Tremor, defined in Supplemental Table S1) were calculated. Demographic data including age and sex of the participants were also collected at the beginning of the study.

### 2.2 Statistical Analysis

The workflow of our proposed comprehensive machine learning based framework is illustrated in Figure 1, which comprises five main steps: 1) data collection and processing; 2) univariate feature screening; 3) univariate association testing; 4) multivariate analysis (using PGEE) to construct a composite digital measure for longitudinal disease progression; 5) performance evaluation. The specifics of each step are described below.

**Figure 1.**
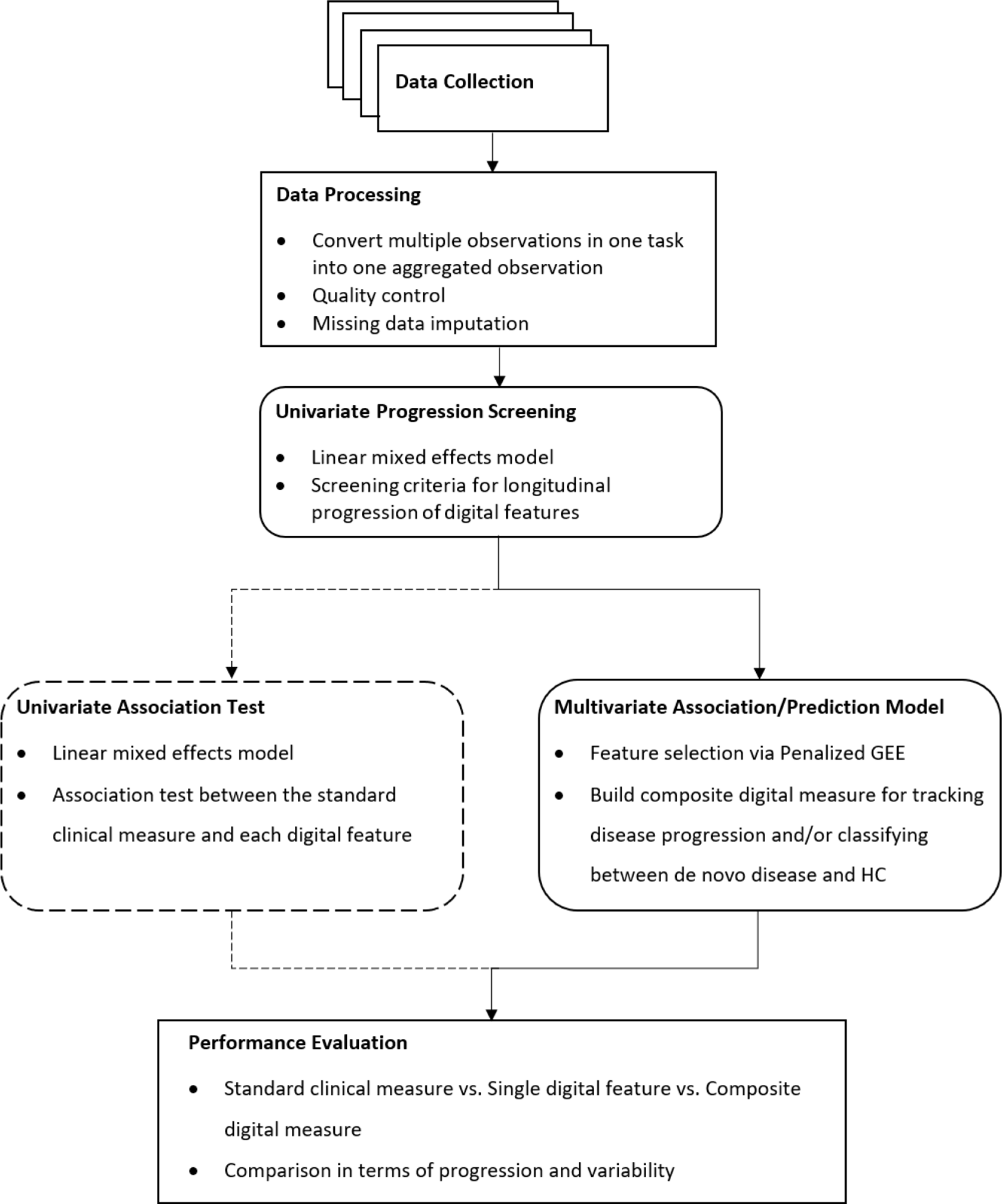
The analysis pipeline to select relevant digital features from high-dimensional DHT data and construct a composite digital measure for disease progression tracking, including 1) DHT data collection and processing, 2) univariate feature progression screening, 3) univariate association test (optional), 4) multivariate/composite digital measure construction, and 5) performance evaluation.

#### 2.2.1 Data Processing and Quality Control

In this first step, data aggregation and pre-processing are performed to convert high-frequency, epoch-level data into a set of aggregated digital features for each task. The movement data collected from DHTs often include epoch-level features (e.g., per second, per minute, or per walking step) that are collected repeatedly during an active task (e.g., two-minute walk). This step simplifies such data and produces a clean, high-dimensional feature set for each participant at each clinical time point, in order to facilitate subsequent longitudinal analyses.

In our illustrative PD example, summary statistics (mean, median, standard deviation, and mean absolution deviation) were calculated to represent the repeated measurements across the entire task for features that had repeated measurements during the task. For example, during the two-minute walk task, step lengths of every step that the participant took were recorded; these were aggregated into task-level features such as mean step length during the two-minute walk task period. After that, we had 256 digital features generated in total. Then, distributions of all features were examined, and the non-informative features that had few distinct values, included a large amount of data missingness, or contained extreme values were removed. For the remaining features, missing data imputation was performed using the mean of available data in each feature. Finally, additional feature quality control steps were implemented, which included removing highly correlated features, log-transforming skewed features, and removing outliers. 141 unique digital features were left for univariate progression screening in the next step.

#### 2.2.2 Univariate Progression Screening

In the second step of our framework, univariate progression screening is recommended to identify whether each digital feature detected disease progression during the study duration. In this step, a linear mixed effects model (LMM) is used to screen the univariate features against a set of pre-determined criteria. Each digital feature is used as the response variable for the screening separately. Independent variables are added to the model as fixed effects, including covariates to be adjusted, group membership, visit, group-by-visit interaction, and covariate-by-visit interactions. Random intercept and slope are added to the model as random effects.

In our illustrative PD example, we applied relatively relaxed screening criteria to select digital features for downstream analysis. We considered a digital feature as a “candidate” if (1) its longitudinal trend was flat in the HC group (i.e., the LMM slope p-value of HC group was larger than 0.05) and (2) it demonstrated a progression trend with time in PD groups (i.e., the LMM group-by-visit interaction p-value was < 0.1 or the p-value of the differential slope between de novo/mild-to-moderate/advanced PD and HC was < 0.1).

#### 2.2.3 Univariate Association Test

To gain additional insights on the univariate associations between the standard clinical measure (i.e., MDS-UPRDS Part III) and the candidate digital features that passed the univariate progression screening, our framework employs a univariate association test step. In this step, a linear mixed effects model is employed, with the clinical measure as the dependent variable and each individual digital feature as the independent variable. Covariates to be adjusted are also included in the model. Random intercepts for each subject are allowed in the model and p-values are calculated to assess the significance of the association between the clinical measure and digital features.

An optional procedure is to further filter the candidate digital features based on their associations to the standard reference measure (i.e., MDS-UPDRS Part III and its subscales in our example) and exclude non-significant features. In our example, we chose to implement relatively relaxed screening criteria to retain more features for the subsequent feature selection, and therefore, we did not exclude features that did not show association with MDS-UPDRS Part III in our downstream analysis.

#### 2.2.4 Multivariate Prediction Model

In the final step of our framework, a multivariate prediction model is developed, which selects and combines a set of digital features that pass the univariate progression screening into a composite digital biomarker of disease progression. As MDS-UPDRS Part III is the current clinical standard for monitoring PD motor function progression in PD clinical trials, this was used as the training endpoint in our illustrative example. As several studies have explored digital measures for disease detection and staging, we proceeded to also include classification between de novo PD and HC as a secondary goal for our composite measure. Importantly, one could use our proposed framework to optimize the measure for disease progression tracking, disease classification, or both, based on the screening criteria and training endpoints used.

To model the high-dimensional longitudinal data, our framework includes a ML-based Penalized Generalized Estimating Equations (PGEE) method [32], which performs simultaneous coefficient estimation and variable selection. Compared to the traditional GEE method, PGEE introduces a penalty term to the estimating function of GEE (details of PGEE is provided in Supplementary Method S1).

To determine the optimal number of digital features (*P*) to be included into the final multivariate prediction model, a cross-validation (CV) strategy is implemented into the framework to avoid overfitting [34]. Specifically, all digital features are first ranked by their PGEE estimates from the training set, then a series of PGEE models with different numbers of top features are built and evaluated in the testing set. The optimal number of features is then determined to be the number of features from the model with the smallest Root Mean Squared Error (RMSE). The approach is further described in Supplementary Method S2.

All candidate digital features that pass the univariate screening are ranked by their PGEE estimates from the largest to the smallest using the whole dataset. The top *P* digital features are used to construct the composite digital measure. Specifically, a GEE model is fitted with the top *P* digital features and covariates as independent variables. The dependent variable is a continuous outcome for the primary goal of tracking disease progression, and a binary outcome for the secondary goal of classifying disease status.

## 3 Results

### 3.1 Patient Demographics and Baseline Characteristics

The baseline demographic characteristics for the participants included in our illustrative analysis are shown in Table 1 and Supplementary Figure S1. The mean ages of four groups (de novo PD, mild-to-moderate PD, advanced PD, and HC) were 66.2, 61.6, 71.2, and 65.6 years, respectively. The ratios of male-to-female subjects in the four groups were 5:5, 9:1, 5:5, and 3:7, respectively.

**Table 1.** Patient baseline characteristics (age and sex) for the three PD groups and healthy control group.

|  | **De novo PD** | **Mild-to-moderate PD (on-therapy)** | **Advanced PD** | **Healthy Control** |
| --- | --- | --- | --- | --- |
| n | 10 | 10 | 10 | 10 |
| Age (years)<br>[mean (SD)] | 66.2 (6.46) | 61.6 (10.76) | 71.2 (4.78) | 65.6 (6.98) |
| Sex<br>[Male (%) / Female (%)] | 5 (50) / 5 (50) | 9 (90) / 1 (10) | 5 (50) / 5 (50) | 3 (30) / 7 (70) |

To determine if age and sex needed be considered covariates to be adjusted for in our models, we calculated the age-by-visit and group-by-visit interaction p-values in linear mixed effects models with MDS-UPDRS Part III as the response. The results, summarized in Supplementary Table S2, suggested that age would affect the slope of MDS-UPDRS Part III progression (with p-value = 0.04) while sex would not (with p-value = 0.19). We therefore considered only age as a covariate to be adjusted in our data analysis models.

### 3.2 Univariate Progression Screening Results

In our illustrative example, our univariate progression screening criteria were such that a digital feature would “pass” if the LMM model for that digital feature showed (1) no progression in the control group and (2) a progression in at least one of the three PD groups. 77 digital features out of 141 screened passed these criteria, including 15 features from postural sway task, 5 features from timed up-and-go (TUG) task, and 57 features from two-minute walk task. Among these, Walk GLLGS (Gait – Lower Limb – Gait Speed) had the smallest group-by-visit interaction p-value (6.0e-07) and the smallest de novo PD vs. HC progression slope p-value (4.7e-04); Walk GLLDS (Gait – Lower Limb – Double Support) had the smallest mild-to-moderate PD vs. HC progression slope p-value (0.01); and Walk GLLSD (Gait – Lower Limb – Step Duration) had the smallest advanced PD vs. HC progression p-value (1.2e-06). P-values of TUG TPV (Timed Up and Go – Turn Peak Velocity) for group-by-visit interaction, de novo PD vs. HC progression slope, mild-to-moderate PD vs. HC progression slope, and advanced PD vs. HC progression slope were 0.008, 0.001, 0.147, and 0.013, respectively. A summary heatmap of all 77 digital features that met the screening criteria is displayed in Figure 2, and the heatmap of all the digital features that were screened is displayed Supplementary Table S3.

**Figure 2.**
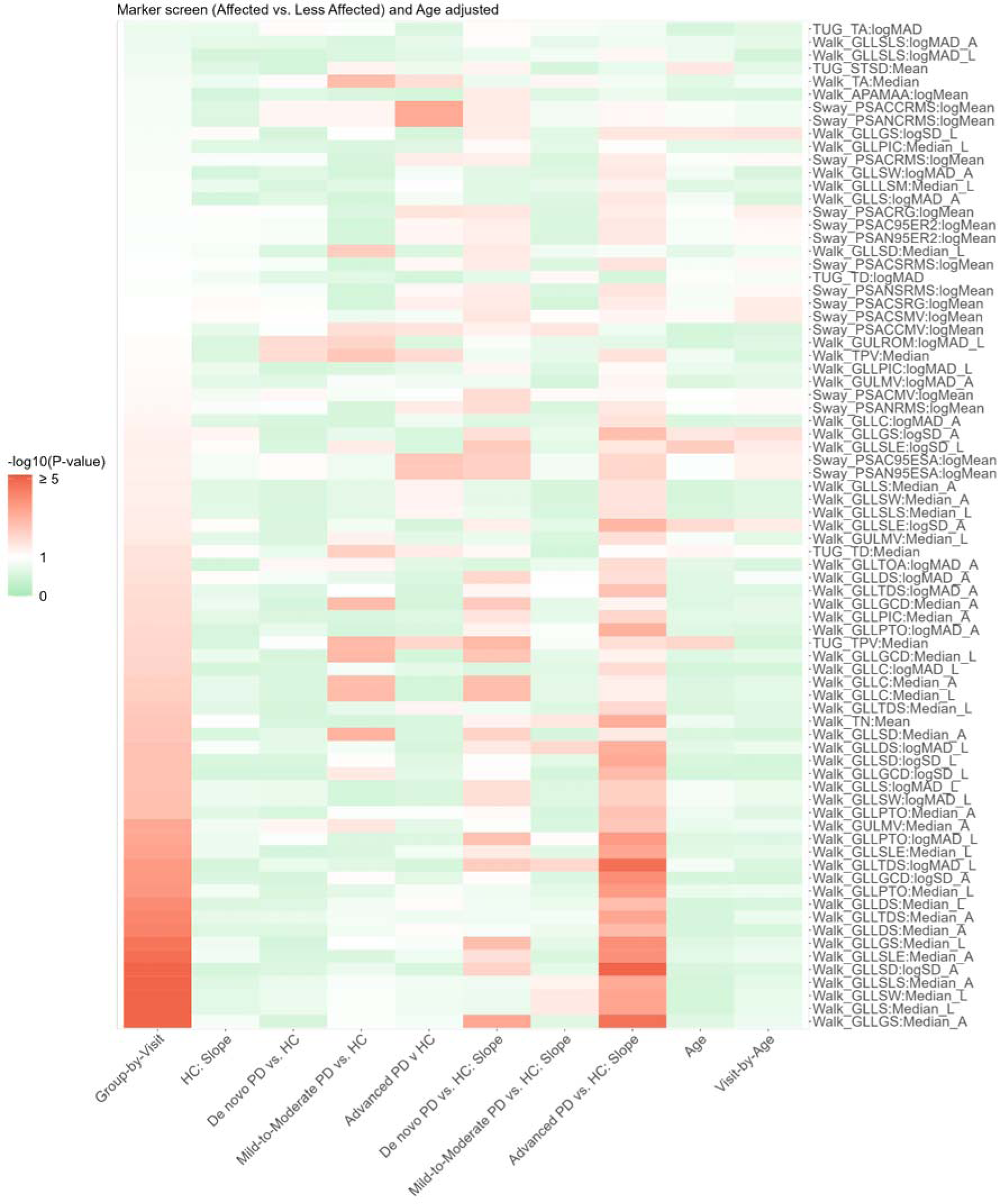
Heatmap representation of the p-values of the 77 digital features that passed the progression screening. The screening criteria applied were (1) no time progression in the HC group (i.e., LMM slope p-value of HC > 0.05), and (2) time progression in at least one of the three PD groups (i.e., LMM group-by-visit interaction p-value < 0.1 or p-value of differential slope between de novo/mild-to-moderate/advanced PD and HC < 0.1).

**Figure 2.**
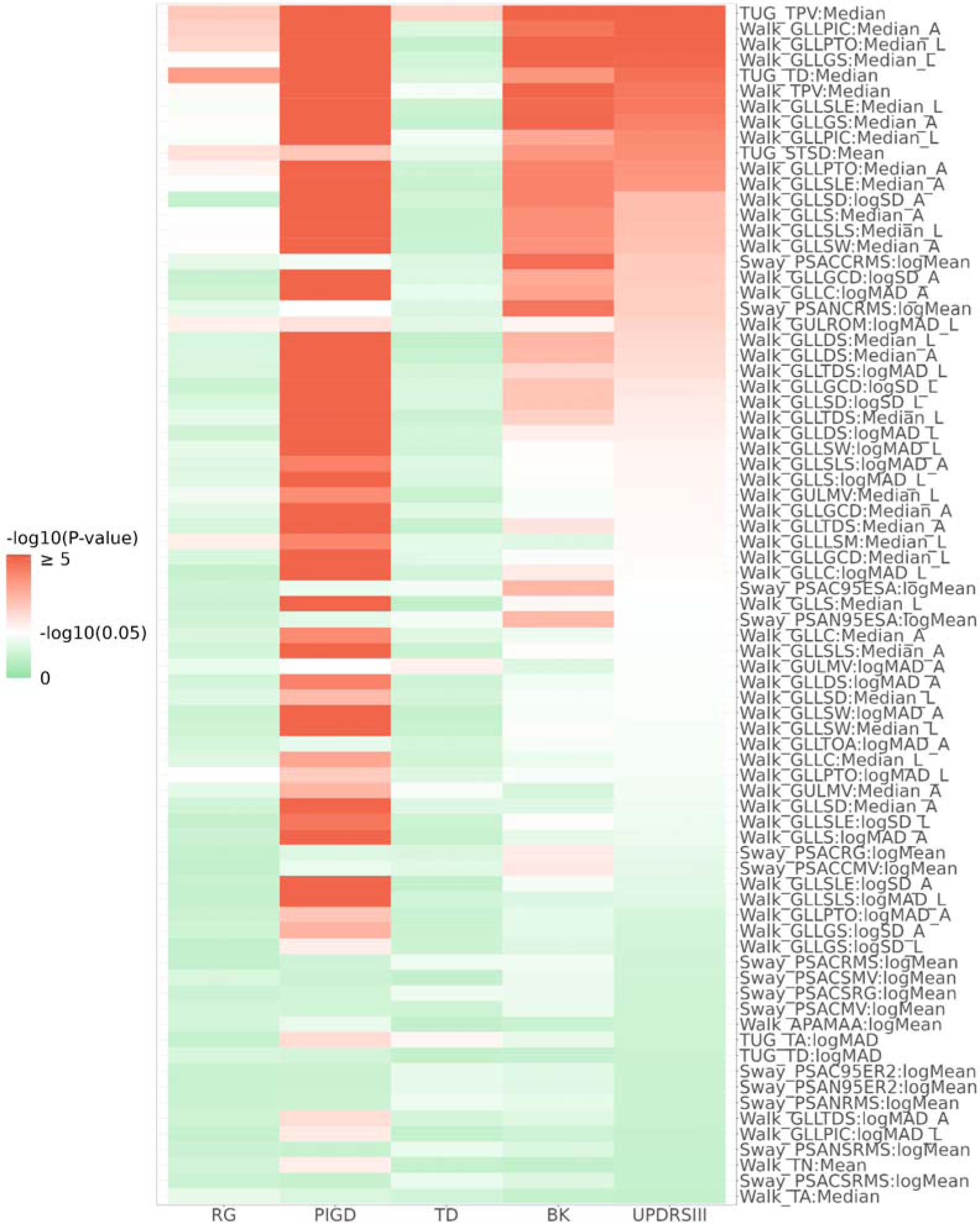
Heatmap of the univariate association testing p-values between MDS-UPDRS Part III (and its subscales: BK, TD, PIGD, RG) and the 77 digital features that passed the univariate screening. P-values were calculated from a linear mixed effects model with MDS-UPDRS Part III or its subscales as the outcome variable. The 77 features were ranked based on their association p-values from the analysis with the MDS-UPDRS Part III score. Each digital feature and age were included as independent variables. Random intercept was added as a random effect.

### 3.3 Univariate Association Analysis Results

Figure 3 shows the univariate association testing results between the 77 digital features that passed the univariate screening in our illustrative example and MDS-UPDRS Part III scores (and its subscales). 37 of these 77 digital features (48.1%) showed significant associations (i.e., p-value < 0.05) with MDS-UPDRS Part III scores (including 32 features from the Walk task, 3 features from the TUG task, and 2 features from the Sway task). The associations of digital features with the MDS-UPDRS Part III scores were generally consistent with their associations with the Bradykinesia (BK) subscale within MDS-UPDRS Part III. Specifically, 40 of the 77 digital features were associated with the BK subscale (including 31 features from the Walk task, 3 features from the TUG task, and 6 features from the Sway task). In addition, 59 of the 77 digital features were associated with the Postural Instability and Gait (PIGD) subscale (including 54 features from the Walk task, 4 features from the TUG task, and 1 feature from the Sway task), while only 3 of the 77 features (TUG TPV, TUG TA, and Walk GULMV) were associated with the Tremor Dominant (TD) subscale.

**Figure 3.**
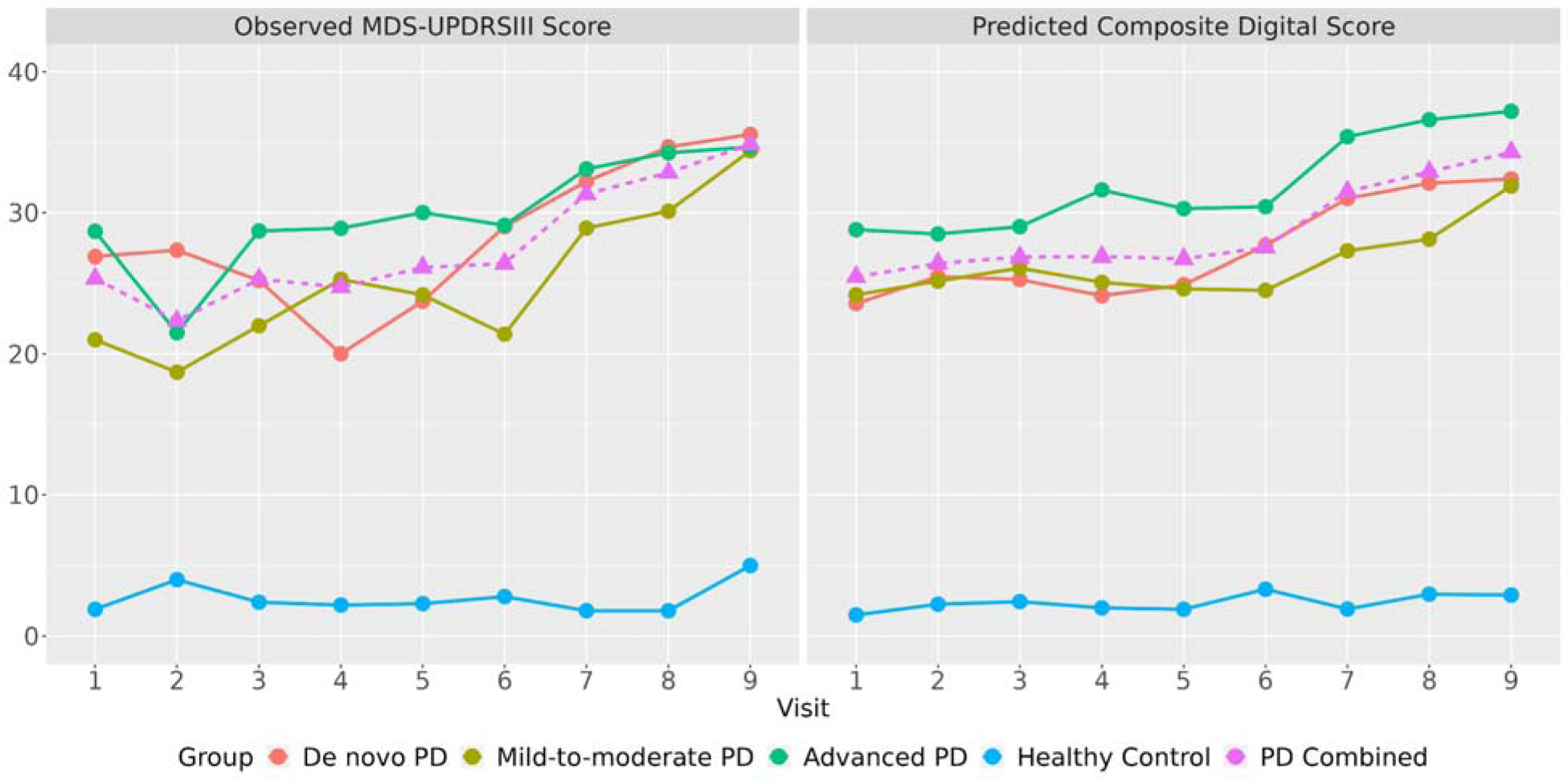
Tracking PD progression via MDS-UPDRS Part III (left panel) and the composite digital measure based on the 11 selected digital features (right panel). The dashed lines represented the observed MDS-UPDRS Part III scores (left panel) and the predicted composite digital scores (right panel) in the three PD groups combined.

Turn Peak Velocity (TPV), obtained from the Timed Up and Go (TUG) test [35], demonstrated the most significant association with MDS-UPDRS Part III. TUG TPV is defined as the maximum achieved angular velocity of trunk rotation in the y-axis during 180-degree turns (deg/sec) and has been found to be related to PD progression in multiple studies [12, 36 – 38]. The progression characteristics of TUG TPV are shown in Figure 4, where the group-wise and subject-wise lines were obtained from the linear mixed effect model and the points represented the observed data. In terms of TUG TPV, the mild-to-moderate, on therapy PD and HC groups were stable, while the de novo and advanced PD groups showed progression.

**Figure 4.**
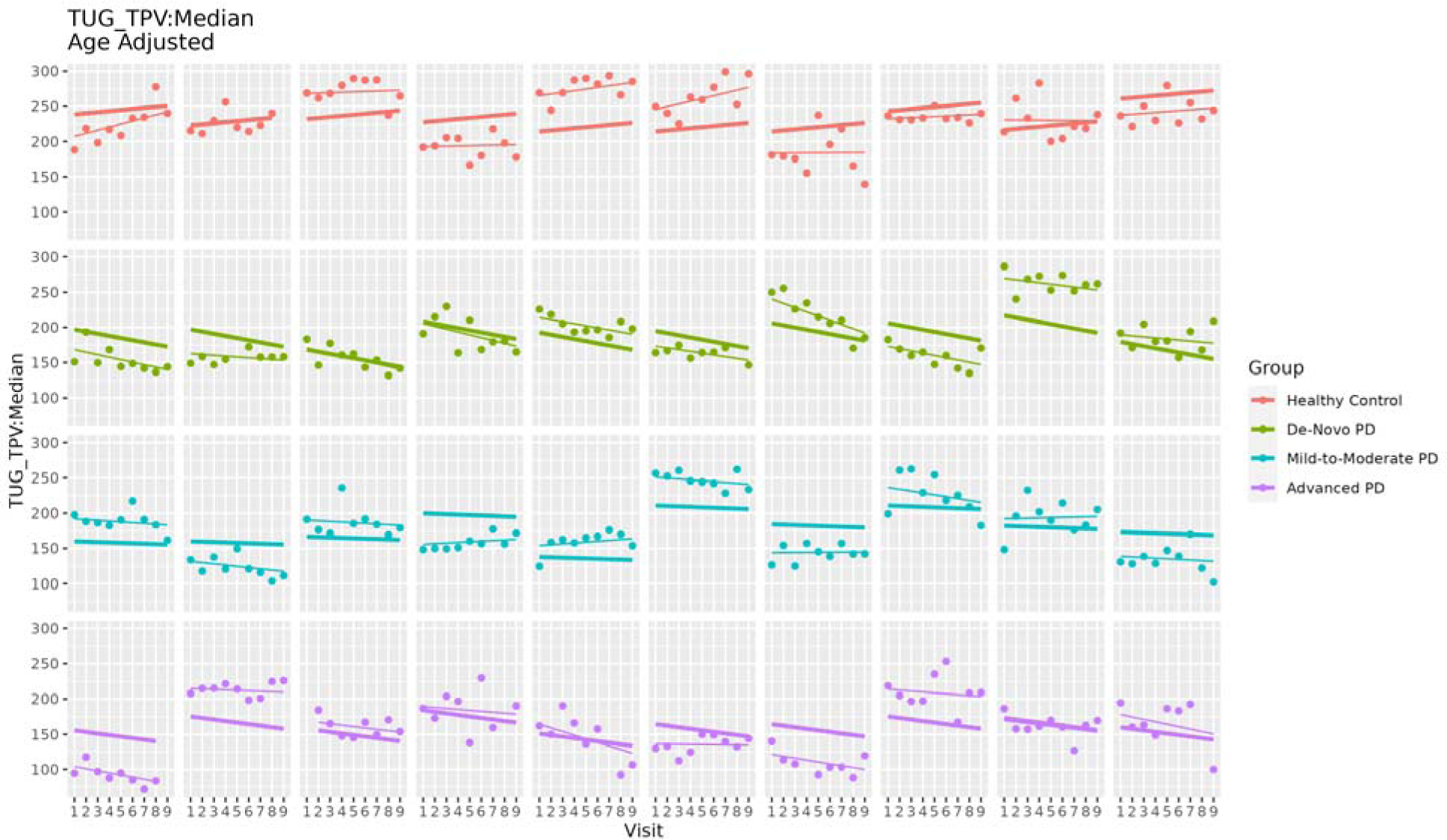
Results from a digital feature, TUG TPV: Turn Peak Velocity (TPV), obtained from the Timed Up and Go (TUG) test, which showed the most significant association with the MDS-UPDRS Part III score. Each row represents the three PD groups and the HC group. Each panel within a row corresponds to a particular subject. The thick lines and thin lines denote the group-wise and subject-wise estimates of progression lines fitted by the linear mixed effects model, respectively. The points denote the observed data.

In general, the univariate association observations were consistent with the progression patterns seen in the MDS-UPDRS Part III and its subscales, which is shown in Supplementary Figure S2. Specifically, compared to the HC group, the BK subscale progressed across all PD groups (at α = 0.1 level). The PIGD subscale progressed in de novo and advanced PD groups while staying stable in the mild-to-moderate, on-therapy PD group. This pattern was similar to most of the digital features included in the analysis, as indicated in Figure 2. In contrast, the TD subscale progressed in the mild-to-moderate, on-therapy PD group, while remaining unchanged in de novo and advanced PD groups.

### 3.4 Multivariate Feature Selection and Prediction Results

#### 3.4.1 Feature Selection

We applied multivariate feature selection to first determine the optimal number of features to be selected for the digital composite measure and subsequently select the digital features for inclusion into the composite score and prediction model in our illustrative example analysis. Supplementary Figure S3 indicated that for the primary analysis goal of developing a composite digital measure for disease progression tracking, using the top 9 features (ranked by their PGEE estimates) overall yielded the smallest RMSE; and for the secondary analysis goal of classifying disease status, using the top 3 features resulted in the largest AUC via internal cross-validation.

We then ranked all pre-screened features (i.e., digital features) according to their PGEE estimates. Nine digital features (TUG TD, TUG TPV, TUG STSD, Walk TA, Walk GLLC, Walk GLLSW, Walk GLLLSM, Walk APAMAA, and Sway PSAN95ESA) were selected for the primary analysis goal; and three digital features (TUG TPV, Walk GLLTOA, and Walk GULMV) were selected for the secondary analysis goal. Table 2 lists the description of these selected features. The selected digital features for both analysis goals were further merged into a digital composite score for the prediction of MDS-UPDRS Part III in the next step. As one of the features (TUG TPV) was selected for both the primary and secondary analysis goals, 11 unique digital features were included in the digital composite score.

**Table 2.** Description of the selected features: 9 for the primary analysis goal of longitudinally tracking MDS-UPDRS Part III score, and 3 for the secondary analysis goal of classifying de novo PD and Healthy Control. MAD = Mean Absolute Deviation, A = Affected side, and L = Less affected side.

| Goal | Feature | Statistic | Side | Description | PGEE Estimate |
| --- | --- | --- | --- | --- | --- |
| Primary | TUG TD | Median |  | Turns - Duration | 0.39 |
|  | TUG TPV | Median |  | Turns - Turn Velocity | -0.38 |
|  | TUG STSD | Mean |  | Stand to Sit - Duration | 0.34 |
|  | Walk TA | Median |  | Turns - Angle | 0.26 |
|  | Walk GLLC | MAD | A | Gait/Lower Limb - Cadence | 0.16 |
|  | Walk GLLSW | MAD | L | Gait/Lower Limb - Swing | 0.07 |
|  | Walk GLLLSM | Median | L | Gait/Lower Limb - Circumduction | -0.06 |
|  | Walk APAMAA | Mean |  | Anticipatory Postural Adjustment - Forward APA Peak | -0.06 |
|  | Sway<br>PSAN95ESA | Mean |  | Postural Sway/Angles - Sway Area | 0.05 |
| Secondary | TUG TPV | Median |  | Turns - Turn Velocity | -0.60 |
|  | Walk GLLTOA | MAD | A | Gait/Lower Limb - Toe Out Angle | -0.43 |
|  | Walk GULMV | Median | A | Gait/Upper Limb - Arm Swing Velocity | -0.27 |

#### 3.4.2 Selected Features for Tracking MDS-UPDRS Part III

The 11 selected unique digital features were merged for the prediction of MDS-UPDRS Part III by fitting a GEE model. The performance was evaluated using the 10-fold CV procedure in PD and HC groups, respectively. As shown in Figure 3, the composite digital measure based on the selected digital features showed a pattern of no change vs. time in the HC group as expected (with RMSE in HC group = 2.8). The composite digital measure had a smoother increasing trend in the overall PD group, as well as each PD subgroup (with RMSE in PD group = 12.7).

We further compared performances among MDS-UPDRS Part III, the composite digital measure, and each of the univariate digital features included in the composite digital measure (e.g., TUG TPV) quantitatively in terms of both progression and variability. Detailed results are summarized in Table 3. Overall, the group-by-visit interaction p-value of composite digital measure was close to that of MDS-UPDRS Part III (7.65e-03 vs. 6.22e-03). The increasing trend of the composite digital measure was much more significant in de novo and advanced PD groups, but didn’t show significant progression in the mild-to-moderate, on-therapy PD group, which was consistent with what is observed in Figure 3. Recall that none of the 11 selected digital features had significant univariate progression in the mild-to-moderate, on-therapy PD group (for example, the mild-to-moderate PD versus HC slope p-value of TUG TPV was not significant, p=0.15). Thus, it was not surprising that the composite digital measure preserved the same pattern. Moreover, the composite digital measure showed smaller between-/within-subject coefficient of variation than MDS-UPDRS Part III. In summary, the results from Figure 3 and Table 3 indicate that the composite digital measure is an attractive aggregated measure for tracking PD progression compared to MDS-UPDRS Part III and to individual digital features.

**Table 3.**
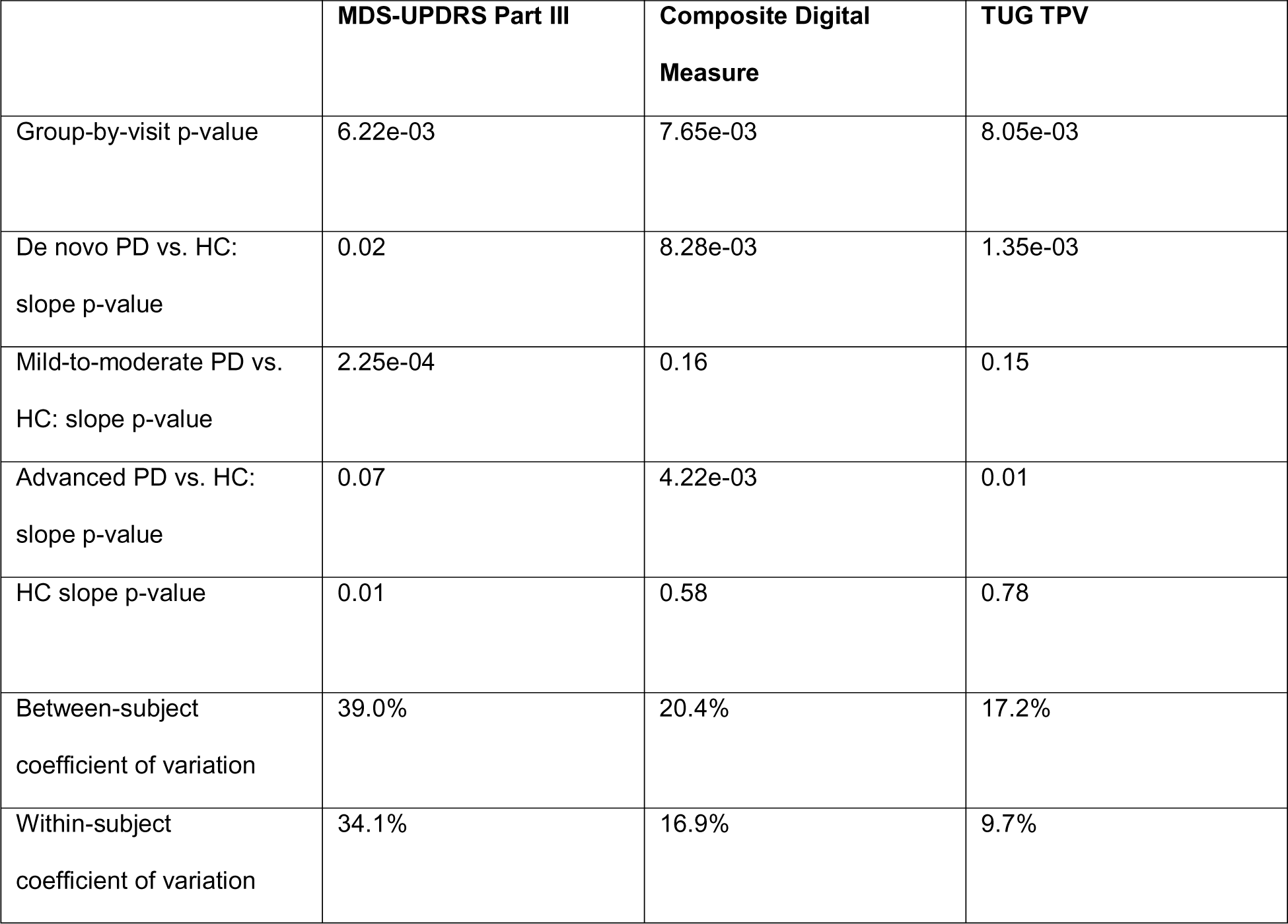
Performance comparison among MDS-UPDRS Part III, the composite digital measure, and TUG TPV in terms of both progression and variability.

|  | **MDS-UPDRS Part III** | **Composite Digital Measure** | **TUG TPV** |
| --- | --- | --- | --- |
| Group-by-visit p-value | 6.22e-03 | 7.65e-03 | 8.05e-03 |
| De novo PD vs. HC:<br>slope p-value | 0.02 | 8.28e-03 | 1.35e-03 |
| Mild-to-moderate PD vs.<br>HC: slope p-value | 2.25e-04 | 0.16 | 0.15 |
| Advanced PD vs. HC:<br>slope p-value | 0.07 | 4.22e-03 | 0.01 |
| HC slope p-value | 0.01 | 0.58 | 0.78 |
| Between-subject<br>coefficient of variation | 39.0% | 20.4% | 17.2% |
| Within-subject<br>coefficient of variation | 34.1% | 16.9% | 9.7% |

#### 3.4.3 Selected Features for Classifying de novo PD and HC

To examine if the composite digital measure developed was also effective in classifying between de novo PD and HC subjects, we trained a GEE classification model using the 11 unique selected digital features (age adjusted). Results are shown in Figure 4. An AUC of 0.81 was achieved, indicating that the composite digital measure (composed of the 11 digital features) had good performance and was effective in classifying between de novo PD and HC subjects.

**Figure 4.**
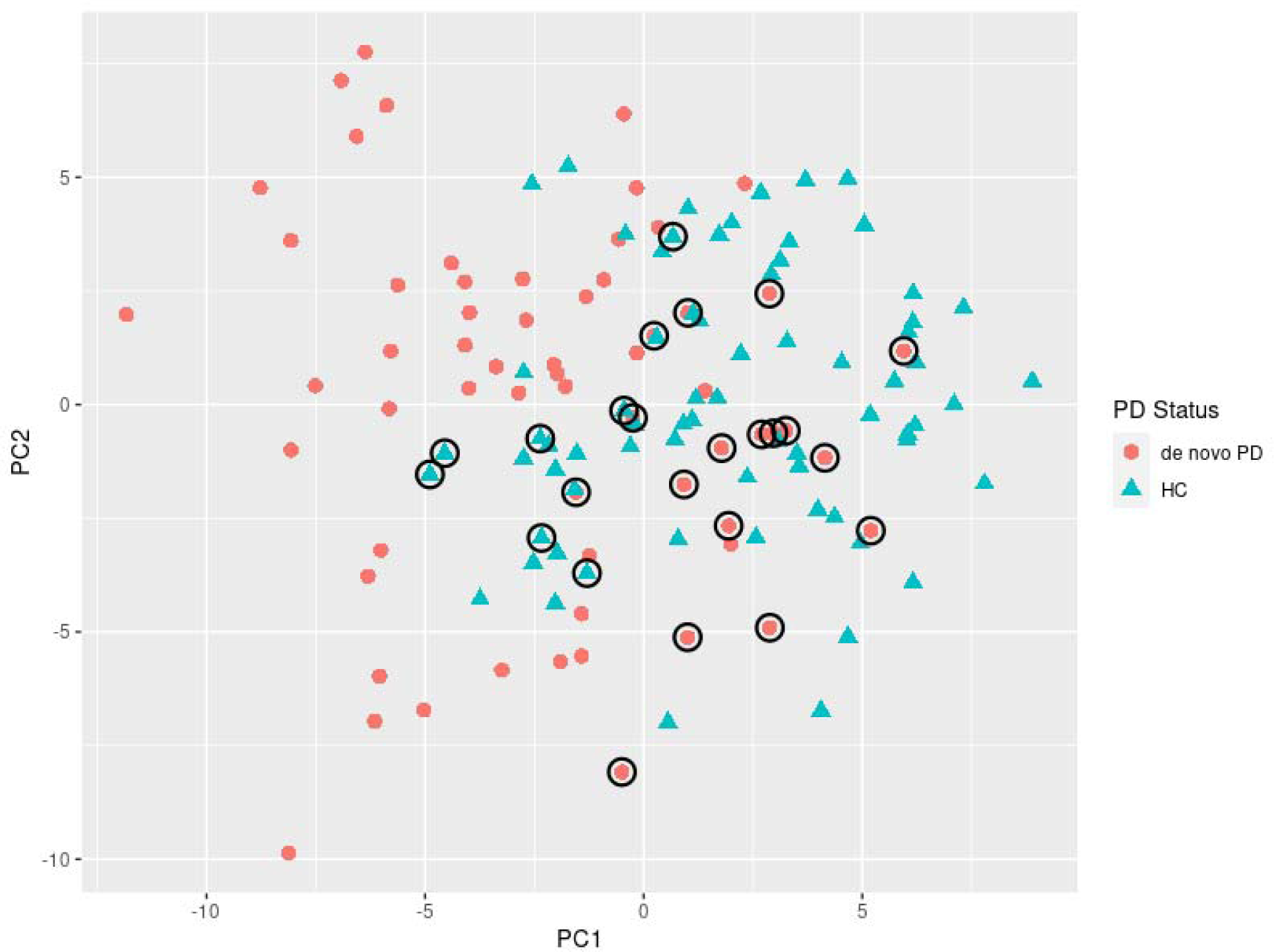
Classification between de novo PD and HC groups using the 11 selected digital features. Each dot indicates the observed PD status (i.e., the ground truth) of each subject at each visit. False classifications (by the prediction model based on 11 selected digital features) are highlighted by black circles.

## 4 Discussion

DHT-derived measures have shown great promise in both tracking disease progression and disease classification. However, it remains challenging to identify digital features for predicting disease progression longitudinally in a high dimensional space, and furthermore, methodologies for combining individual digital features into composite digital measures have not been standardized. Features derived from DHT data may be high dimensional, with various data types and complex correlation structures. In this paper, we propose a principled and comprehensive methodology for the identification of relevant digital features of disease progression from large DHT data sets, and subsequent construction of a composite digital measure for disease progression tracking. Specifically, in Step 1, data is collected and processed for aggregated observation and quality control. In Step 2, we apply a linear mixed effects model for univariate screening for longitudinal progression of digital features. In Step 3, a univariate association test is conducted between candidate digital features (i.e., features that pass the univariate screening) and clinical scores, for example the MDS-UPDRS Part III and/or its subscales. In Step 4, the candidate digital features are ranked via a ML-based method, PGEE, for high-dimensional longitudinal data analysis. The optimal number of top features to be included into the composite digital measure is further determined using a cross-validation based algorithm to avoid overfitting.

To demonstrate the utility of our methodology, we applied it to data collected from a PD longitudinal observational study, which consisted of Opal^TM^ sensor-based movement measurements and MDS-UPDRS Part III scores collected from PD patients at a range of disease stages and healthy controls over a 2-year duration. The composite digital measure we developed generally showed a smoother and more significant increasing trend in PD groups and smaller between-/within-subject coefficients of variation than MDS-UPDRS Part III in this small dataset (N=40), indicating potential utility for the composite digital measures to be used to track disease progression more sensitively and with less variability vs. standard clinical measures. It should be noted that the dataset in our illustrative examples was small (N=40), and therefore, results of our analysis should be interpreted with caution. The analysis reported here was presented as an illustrative example of our proposed methodology and framework and not intended as a proposed composite measure for use in future studies. We also note that the composite digital measure shows less significant progression trending in mild-to-moderate, on-therapy PD patients compared to in de novo and advanced PD patients. This outcome is consistent with the trends observed by Brzezicki et al. [11] using data derived from the OxQUIP study. We further evaluated performances of the composite digital measure built from our methodology for classifying between de novo PD patients and HC. The measure had an AUC ROC of 0.81, indicating that the composite digital measure (composed of the 11 digital features) had good performance and was effective in classifying between de novo PD and HC subjects.

Note that in our analysis, the top digital features (i.e., those with the largest PGEE estimates from the multivariate penalized regression model) were selected for both the primary analysis goal of tracking MDS-UPDRS Part III progression and the secondary analysis goal of classifying between de novo PD and HC. While the digital feature TUG TPV ranked high in both subgroups of selected features, we observe that the digital features that reflect disease progression are not necessarily the same as digital features that reflect classification. The final composite digital measure constructed with the merged features, on one hand, keeps the main characteristics of single digital features (i.e., progressing in de novo and advanced PD groups, but being flat in mild-to-moderate, on-therapy PD and HC groups). On the other hand, the composite digital measure has a more significant increasing longitudinal trend compared to single digital features (including TUG TPV). This finding is not unique to composite digital measures as many traditional clinical measures (e.g., MDS-UPDRS Part III) are composite in nature and exhibit this trend. It is worth noting that the composite digital measure constructed following our proposed pipeline is comprised of features with diverse measurement properties. It is not a combination of the best-performing individual features (i.e., neither features with the most progressions in PD groups nor features with the most significant univariate association with MDS-UPDRS Part III). A possible explanation is that combining top features with high correlations doesn’t necessarily add additional information to the composite score; there could be redundancy among digital features. It also suggests opportunities to further improve the performance of the composite digital measure by enriching the feature set with different assessments/tasks and measures.

The superior performance observed in the multivariate analysis, albeit from a small pilot dataset, suggests promise for use of composite digital measures for progression tracking in future studies. Recent modelling efforts have shown that an increased precision made possible by more objective and frequent composite digital measures could enable smaller and shorter proof-of-concept studies to demonstrate disease-modifying treatment effect [20]. Open discussions on methodologies to identify the relevant digital features (from the multitude of digital measure possible with DHTs) and construct composite digital measures are critical to enable construction and use of such digital measures, and we present a methodology for this herein.

There are several limitations of our work. First, a major caveat of the results reported from the illustrative example herein is that this analysis only used a small number of participants. Our proposed analysis workflow for digital biomarker development needs to be applied to additional studies with larger N to further demonstrate utility. The identified individual digital features of Parkinson’s disease progression and the composite digital measure presented herein is solely for purposes of illustrating the methodology approach. They would need to be validated and verified in an independent dataset in further research before they can be used as digital biomarkers of disease progression and treatment response. Second, the digital features in our study were obtained from sensor-based movement measurements using one DHT system used during supervised, in-clinic tasks. Different or expanded digital features may be available with different DHTs, different task-based assessments, use of passive monitoring approaches, technology evolution, and further algorithm development. It is worth noting that we mainly use this feature set to demonstrate the methodology, and our proposed high-dimensional longitudinal data analysis framework (including feature selection and predictive modelling) is adaptive for different feature sets collected from different sensor technologies.

Notably, the current dataset is longitudinal but only contains in-clinic visit data. One advantage of DHTs is that they may offer the ability to capture data outside of the clinic much more frequently. Other studies, including the Phase 2 Trial of Anti α-Synuclein Antibody in Early Parkinson’s Disease (PASADENA) study [10] (daily tasks) and the Personalized Parkinson Project (PPP) study [39] (bi-weekly tasks), have shown utility in capturing remotely collected DHT data with increased measurement frequency. Increased measurement frequency could further enhance the performance of digital measures in quantifying disease progression, as it could address the day-to-day symptom fluctuations and reduce the measurement variability. Such remotely acquired digital features could also be applied to the methodology and framework we’ve reported here.

In addition, there is emerging research into characterization of the neurodegenerative disease progression directly from raw sensor signals recorded by DHTs (e.g., wearable sensors, environmental sensors, smartphone sensors) using deep neural networks and other black box algorithms [40, 41]. Germane to these efforts is an important question about the interpretability of the ensuing models and results [42, 43]. In our work, we identified candidate digital features of disease progression using inherently interpretable linear models. We did not explore deep learning of the raw sensor data directly; such an approach is an interesting future direction of research.

In summary, with the rapid development of DHTs, digital measures are playing an increasingly important role in not only neurodegenerative disease detection, but also longitudinally tracking disease progression over time and detection of therapeutic response. Our proposed ML-based framework for identifying digital features of progression and constructing composite digital measures adds to the existing body of literature on digital measure analysis methodologies and may help accelerate the translation of digital measures to utility for drug development and clinical practice.

## Data Availability

The original data presented in this paper is from the ongoing OxQUIP study and cannot be shared until completion of the whole study and full dissemination of results. This is expected to become possible within 24 months from the end of the study. Qualified researchers will be able to contact the Principal Investigator at the University of Oxford.

## References

1. Goetz, C.G., Tilley, B.C., Shaftman, S.R., et al. 2008. Movement Disorder Society-sponsored revision of the Unified Parkinson’s Disease Rating Scale (MDS-UPDRS): scale presentation and clinimetric testing results. Movement disorders: official journal of the Movement Disorder Society 23 (15):2129–2170.

2. Committee for Medicinal Products for Human Use. 2018. Qualification opinion on dopamine transporter imaging as an enrichment biomarker for Parkinson’s disease clinical trials in patients with early Parkinsonian symptoms. European Medicines Agency:EMA/CHMP/SAWP/765041/2017.

3. Horak, F.B. and Mancini, M. 2013. Objective features of balance and gait for Parkinson’s disease using body-worn sensors. Movement Disorders 28 (11):1544–1551.

4. Heldman, D.A., Espay, A.J., LeWitt, P.A., et al. 2014. Clinician versus machine: reliability and responsiveness of motor endpoints in Parkinson’s disease. Parkinsonism & related disorders 20 (6):590–595.

5. LeBaron, V., Hayes, J., Gordon, K., et al. 2019. Leveraging smart health technology to empower patients and family caregivers in managing cancer pain: protocol for a feasibility study. JMIR Research Protocols 8 (12):e16178.

6. Evers, L.J., Raykov, Y.P., Krijthe, J.H., et al. 2020. Real-life gait performance as a digital feature for motor fluctuations: the Parkinson@ Home validation study. Journal of medical Internet research 22 (10):e19068.

7. Erb, M.K., Karlin, D.R., Ho, B.K., et al. 2020. mHealth and wearable technology should replace motor diaries to track motor fluctuations in Parkinson’s disease. NPJ digital medicine 3 (1):6.

8. Mahadevan, N., Demanuele, C., Zhang, H., et al. 2020. Development of digital features for resting tremor and bradykinesia using a wrist-worn wearable device. NPJ digital medicine 3 (1):5.

9. Burq, M., Rainaldi, E., Ho, K.C., et al. 2022. Virtual exam for Parkinson’s disease enables frequent and reliable remote measurements of motor function. NPJ Digital Medicine 5 (1):65.

10. Pagano, G., Boess, F.G., Taylor, K.I., et al. 2021. A phase II study to evaluate the safety and efficacy of prasinezumab in early Parkinson’s disease (PASADENA): rationale, design, and baseline data. Frontiers in Neurology 12:705407.

11. Brzezicki, M.A., Conway, N., Sotirakis C., et al. 2023. Antiparkinsonian medication masks motor signal progression in de novo patients. Heliyon:e16415.

12. Pagano, G., Taylor, K.I., Anzures-Cabrera, J., et al. 2022. Trial of prasinezumab in early-stage Parkinson’s disease. New England Journal of Medicine 387 (5):421–432.

13. Robin, J., Xu, M., Detke, M., et al. 2022. Validation of an objective, speech-based object content score for measuring disease progression in AD. https://winterlightlabs.com/assets/publications/ctad-2022-longitudinal_ad_objectscore_final.pdf.

14. Liu, Y., Zhang, G., Tarolli, C.G., et al. 2022. Monitoring gait at home with radio waves in Parkinson’s disease: A marker of severity, progression, and medication response. Science Translational Medicine 14 (663):eadc9669.

15. Yang, Y., Yuan, Y., Zhang, G., et al. 2022. Artificial intelligence-enabled detection and assessment of Parkinson’s disease using nocturnal breathing signals. Nature medicine 28 (10):2207–2215.

16. Kadirvelu, B., Gavriel, C., Nageshwaran, S., et al. 2023. A wearable motion capture suit and machine learning predict disease progression in Friedreich’s ataxia. Nature Medicine:1–9.

17. Ricotti, V., Kadirvelu, B., Selby, V., et al. 2023. Wearable full-body motion tracking of activities of daily living predicts disease trajectory in Duchenne muscular dystrophy. Nature Medicine:1–9.

18. Johnson, S.A., Karas, M., Burke, K.M., et al. 2023. Wearable device and smartphone data quantify ALS progression and may provide novel outcome measures. NPJ Digital Medicine 6(1):34.

19. Giboin L.S., Simillion, C., Rennig, J., et al. 2023. A digital motor score for sensitive detection of disease progression in early manifest Huntington’s disease. https://medically.roche.com/global/en/medical-material.a86d594d-0db2-42d9-9bef-6622e0b574d2.qr.html?cid=slprxx2304nehdchdi2023.

20. Mori, H., Wiklund, S.J. and Zhang, J.Y. 2022. Quantifying the Benefits of Digital Features and Technology-Based Study Endpoints in Clinical Trials: Project Moneyball. Digital Features 6 (2):36–46.

21. Adams, J.L., Kangarloo, T., Tracey, B., et al. 2023. Using a smartwatch and smartphone to assess early Parkinson’s disease in the WATCH-PD study. npj Parkinson’s Disease 9(1):64.

22. Perumal, S.V. and Sankar, R. 2016. Gait and tremor assessment for patients with Parkinson’s disease using wearable sensors. Ict Express 2(4):168–174.

23. Dinov, I.D., Heavner, B., Tang, M., et al. 2016. Predictive big data analytics: a study of Parkinson’s disease using large, complex, heterogeneous, incongruent, multi-source and incomplete observations. PloS one 11 (8):e0157077.

24. Alaskar, H. and Hussain, A. 2018. Prediction of Parkinson disease using gait signals. In 2018 11th international conference on developments in eSystems engineering (DeSE):23-26.

25. Gao, C., Sun, H., Wang, T., et al. 2018. Model-based and model-free machine learning techniques for diagnostic prediction and classification of clinical outcomes in Parkinson’s disease. Scientific reports 8 (1):7129.

26. Tsoulos, I.G., Mitsi, G., Stavrakoudis, A., et al. 2019. Application of machine learning in a parkinson’s disease digital feature dataset using Neural Network Construction (NNC) methodology discriminates patient motor status. Frontiers in ICT 6:10.

27. Dadu, A., Satone, V., Kaur, R., et al. 2022. Identification and prediction of Parkinson’s disease subtypes and progression using machine learning in two cohorts. NPJ Parkinson’s Disease 8 (1):172.

28. Czech, M.D., Badley, D., Yang, L., et al. 2024. Improved measurement of disease progression in people living with early Parkinson’s disease using digital health technologies. Communications Medicine 4(1):49.

29. Taylor, K., Lipsmeier, F., Scelsi, M., et al. 2024. Exploratory Sensor-based Outcome Measures Show Divergent Slopes of Motor Sign Progression in Parkinson’s Disease Patients Treated with Prasinezumab. Preprint at 10.21203/rs.3.rs-3921378/v1.

30. Sotirakis, C., Su, Z., Brzezicki, M.A., et al. 2023. Identification of motor progression in Parkinson’s disease using wearable sensors and machine learning. npj Parkinson’s Disease 9 (1):142.

31. Liang, K.Y. and Zeger, S.L. 1986. Longitudinal data analysis using generalized linear models. Biometrika 73 (1):13–22.

32. Wang, L., Zhou, J. and Qu, A. 2012. Penalized generalized estimating equations for high-dimensional longitudinal data analysis. Biometrics 68 (2):353–360.

33. Pereira, M.F., Buchanan, T., Höglinger, G.U., et al. 2022. Longitudinal changes of early motor and cognitive symptoms in progressive supranuclear palsy: the OxQUIP study. BMJ Neurology Open 4:1.

34. Svetnik, V., Liaw, A., Tong, C., et al. 2004. Application of Breiman’s random forest to modeling structure-activity relationships of pharmaceutical molecules. In Multiple Classifier Systems: 5th International Workshop (MCS) 5:334-343.

35. Zampieri, C., Salarian, A., Carlson-Kuhta, P., et al. 2010. The instrumented timed up and go test: potential outcome measure for disease modifying therapies in Parkinson’s disease. *Journal of Neurology*, Neurosurgery & Psychiatry 81 (2):171–176.

36. Koop, M.M., Ozinga, S.J., Rosenfeldt, A.B., et al. 2018. Quantifying turning behavior and gait in Parkinson’s disease using mobile technology. IBRO reports 5:10–16.

37. Lowry, K., Woods, T., Malone, A., et al. 2022. The Figure-of-8 Walk Test used to detect the loss of motor skill in walking among persons with Parkinson’s disease. Physiotherapy Theory and Practice 38 (4):552–560.

38. Lipsmeier, F., Taylor, K.I., Postuma, R.B., et al. 2022. Reliability and validity of the Roche PD Mobile Application for remote monitoring of early Parkinson’s disease. Scientific reports 12 (1):1–15.

39. Bloem, B.R., Marks, W.J., Silva de Lima, A.L., et al. 2019. The Personalized Parkinson Project: examining disease progression through broad features in early Parkinson’s disease. BMC neurology 19 (1):1–10.

40. Atri, R., Urban, K., Marebwa, B., et al. 2022. Deep Learning for Daily Monitoring of Parkinson’s Disease Outside the Clinic Using Wearable Sensors. Sensors 22 (18):6831.

41. Khan, P., Kader, M.F., Islam, S.R., et al. 2021. Machine learning and deep learning approaches for brain disease diagnosis: principles and recent advances. IEEE Access 9:37622–37655.

42. Rudin, C., Chen, C., Chen, Z., et al. 2022. Interpretable machine learning: Fundamental principles and 10 grand challenges. Statistic Surveys 16:1–85.

43. US Food and Drug Administration 2021. Good machine learning practice for medical device development: Guiding principles. https://www.fda.gov/medical-devices/software-medical-device-samd/good-machine-learning-practice-medical-device-development-guiding-principles.

