## Supplementary Materials for "A Novel Machine Learning Based Framework for Developing Composite Digital Biomarkers of Disease Progression"

Supplementary Method S1 – S2

Supplementary Figure S1 – S3

Supplementary Table S1 – S3

### Supplementary Methods

**Method S1. Penalized Generalized Estimating Equations (PGEE) for high-dimensional longitudinal data analysis**

The Generalized Estimating Equations (GEE) approach has been widely applied to longitudinal data analysis. However, the direct use of traditional GEE in high-dimensional longitudinal data may lead to misleading results. In this analysis, we chose to use a ML-based Penalized GEE (PGEE) method, which performs simultaneous coefficient estimation and variable selection for longitudinal data analysis with high-dimensional variables (i.e., digital features). Specifically, the estimating function in GEE is defined as

$$S\left( \beta\right)=\frac{1}{N}\sum_{i=1}^{N} X_{i}^{T}V_{i}^{-1}\left( \beta\right)\left( Y_{i}-\mu_{i}\left( \beta\right) \right)=0, where V_{i}\left( \beta\right)=A_{i}^{{-1}/2}\left( \beta\right) R A_{i}^{1/2}\left( \beta\right).$$

Here, $\beta=\left( \beta_{1},\cdots,\beta_{p} \right)$ is the regression coefficients, and $p$ is the number of digital features. $N$ is the total number of subjects. $R$ is a working correlation matrix. $X_{i}=\left( X_{i1},\cdots,X_{in_{i}} \right)^{T}$ denotes the $n_{i}\times p$ matrix of covariates, and $n_{i}$ is the number of visits for subject $i$. $A_{i}\left( \beta\right)$ is an $n_{i}\times n_{i}$ diagonal matrix with the marginal variance of responses.

We further add a penalty term to $S\left( \beta\right)$ in the PGEE estimating function:

$$U\left( \beta\right)=S\left( \beta\right)-q_{\lambda}\left( \left| \beta\right| \right)\circ sign\left( \beta\right), where q_{\lambda}\left( \left| \beta\right| \right)=\left( q_{\lambda}\left( \left| \beta_{1} \right| \right),\cdots,q_{\lambda}\left( \left| \beta_{P} \right| \right) \right)^{T},$$

$$q_{\lambda}\left( t \right)=\lambda\times I\left( t<\lambda\right)+\frac{a\lambda-t}{a-1}\times I\left( \lambda\leq t<a\lambda\right)+0\times I\left( t\geq a\lambda\right).$$

Here, the tuning parameter $\lambda$ determines the degree of shrinkage. Following the suggestion of Fan and Li (2001), we set $a=3.7$. The notation $\circ$ denotes the component-wise product. And $q_{\lambda}\left( t \right)$ is the penalty function. Specifically, if $t<\lambda$, then $q_{\lambda}\left( t \right)=\lambda$, indicating that the GEE $S\left( \beta\right)$ is fully penalized; on the other hand, if $t\geq a\lambda$, then $q_{\lambda}\left( t \right)=0$, suggesting that the GEE $S\left( \beta\right)$ is not penalized.

**Method S2. Algorithm of determining the optimal number of top features**

| **Algorithm 1: Determine the optimal number of top features via cross-validation** | |
| --- | --- |
| **Hyper-parameter:** $P$  **Initialization:** Working correlation matrix $R$, penalty $\lambda$, $K$-fold CV | |
| **while** $i\leq K$ **do** | |
|  | Rank all features (i.e., digital features) in terms of their PGEE estimates from the training set. |
|  | Train different PGEE models with different number of top features ($P$) in the training set. |
|  | Apply the PGEE models to the testing set, record the predicted outcomes of subjects in the testing set. |
| **end** | |
| Calculate Root Mean Square Error (RMSE) or Area under the Curve (AUC) across all subjects with their observed and predicted outcomes, depending on the type of the endpoint. The optimal number of top features ($P$) is determined as the one with the smallest RMSE for the disease progression endpoint or the largest AUC for the disease status endpoint. | |
| **Output:** $P$, RMSE or AUC | |

### Supplementary Figures

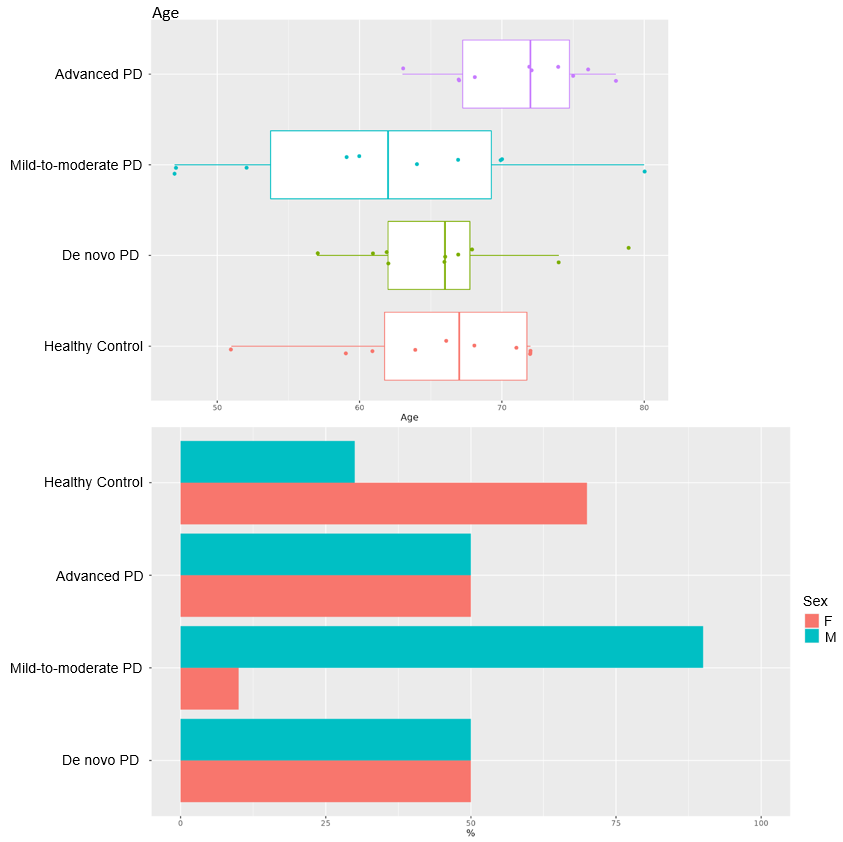

**Figure S1.** Distributions of age (top) and sex (bottom) in four different groups: de novo PD, mild-to-moderate, on-therapy PD, advanced PD, and HC.

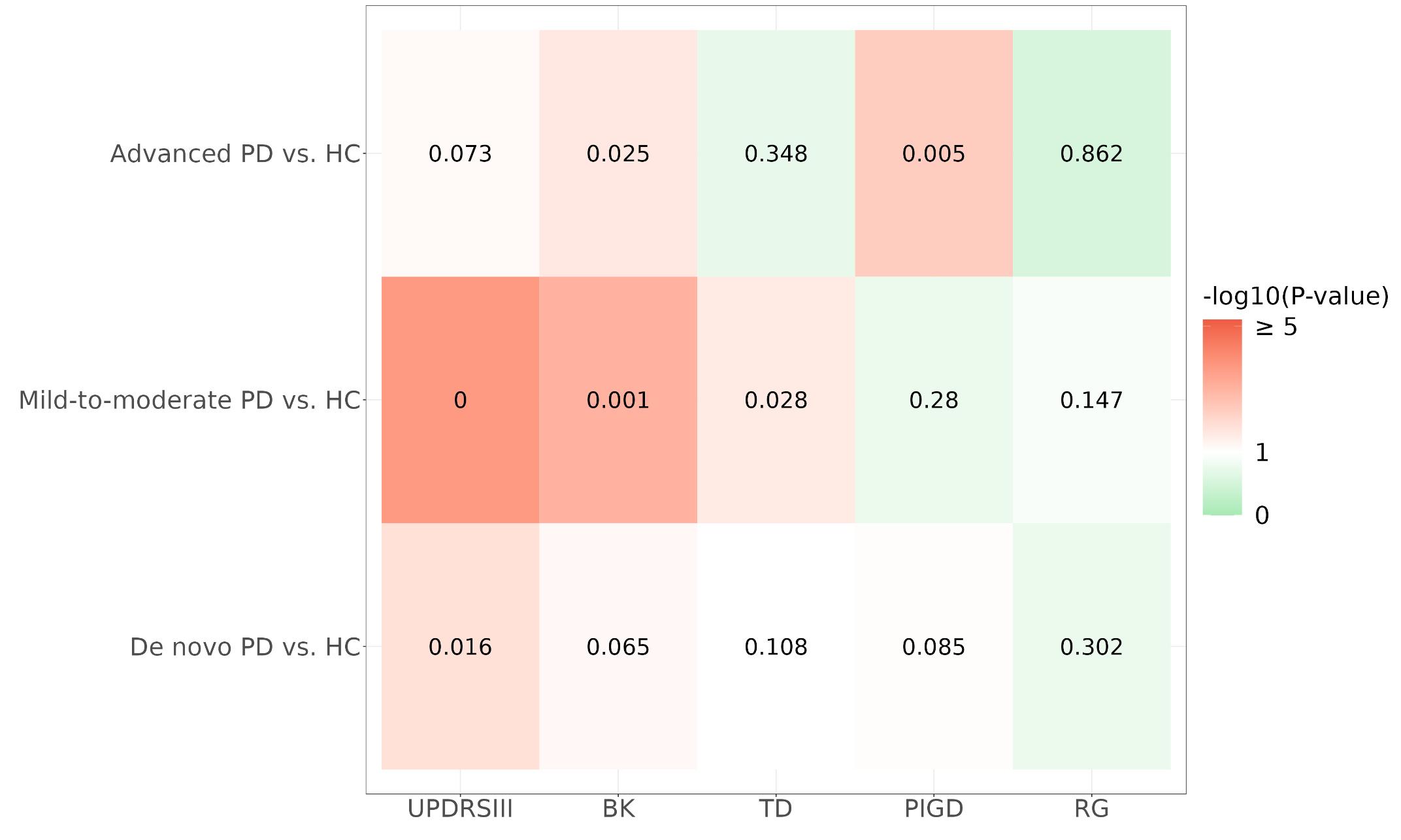

**Figure S2.** Heatmap of differential slope (i.e., progression) p-values between PD groups (i.e., de novo PD, mild-to-moderate PD, and advanced PD) and HC group using MDS-UPDRS Part III total score and its domain sub-scores. P-values were calculated from a linear mixed effects model with UPDRS III or its subscales (i.e., BK, TD, PIGD, and RG) as outcome variable. The independent variables included as fixed effects were intercept, group membership, age, visit, group-by-visit interaction, and age-by-visit interaction. Random intercept and slope were added as random effects. Original p-values were shown in the plot (with p-value threshold $\alpha$ = 0.1).

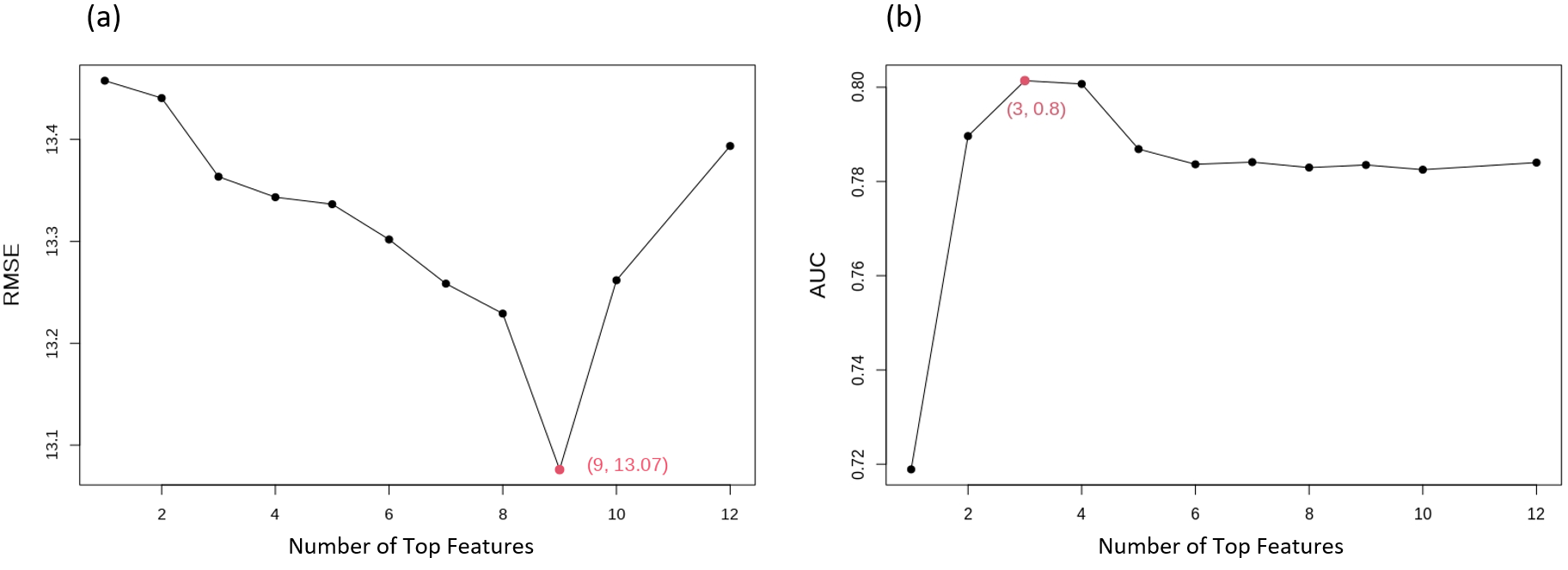

**Figure S3.** The optimal number of top features (i.e., digital features) to be selected into (a) MDS-UPDRS Part III prediction model, and (b) de novo PD versus HC classification model via a cross-validation procedure.

### Supplementary Tables

**Table S1.** MDS-UPDRS Part III Subscales used in the analysis.

| **UPDRS Part III Subscale** | **Items from MDS-UPDRS Part III** |
| --- | --- |
| Bradykinesia (BK) | 3.13 Posture  3.14 Global spontaneity of movement (body bradykinesia)  3.4a Finger tapping LEFT HAND  3.4b Finger tapping RIGHT HAND  3.5a Hand movements LEFT HAND  3.5b Hand movements RIGHT HAND  3.6a Pronation supination LEFT HAND  3.6b Pronation supination RIGHT HAND  3.7a Toe tapping LEFT FOOT  3.7b Toe tapping RIGHT FOOT  3.8a Leg agility LEFT FOOT  3.8b Leg agility RIGHT FOOT  3.9 Arising from chair |
| Rigidity (RG) | 3.3a Rigidity in NECK  3.3b Rigidity in LEFT ARM  3.3c Rigidity in RIGHT ARM  3.3d Rigidity in LEFT LEG  3.3e Rigidity in RIGHT LEG |
| Postural Instability and Gait Difficulty (PIGD) | 3.10 Gait  3.11 Freezing of gait  3.12 Postural stability |
| Tremor dominant (TD) | 3.15a Postural tremor LEFT HAND  3.15b Postural tremor RIGHT HAND  3.16a Kinetic tremor LEFT HAND  3.16b Kinetic tremor RIGHT HAND  3.17a Rest tremor LEFT ARM  3.17b Rest tremor LEFT LEG  3.17c Rest tremor RIGHT ARM  3.17d Rest tremor RIGHT LEG  3.17e Rest tremor LIP AND JAW  3.18 Constancy of rest tremor |

**Table S2.** Determining whether sex and age (as covariates) would affect UPDRS III level at baseline and progression over time.

| A. lme(UPDRS III $\sim$ Sex*Visit, random = $\sim$Visit\|Subject ID, data = PD patients) | | | |
| --- | --- | --- | --- |
| Covariate | Estimate | SE | P-value |
| Sex | 0.83 | 4.46 | 0.85 |
| Visit | 1.66 | 0.29 | 0.01 |
| Sex:Visit | -0.66 | 0.50 | 0.19 |
| B. lme(UPDRS III $\sim$ Age*Visit, random = $\sim$Visit\|Subject ID, data = PD patients) | | | |
| Covariate | Estimate | SE | P-value |
| Age | 0.26 | 0.24 | 0.30 |
| Visit | -2.11 | 1.79 | 0.24 |
| Age:Visit | 0.05 | 0.03 | 0.04 |

**Table S3.** 77 Digital features that passed the univariate progression screening, including 15 postural sway features, 5 timed up-and-go features, and 57 two-minute walk features. P-values were calculated using linear mixed effects models.

| Feature | P-value: group-by-visit | P-value: slope of de novo PD vs. HC | P-value: slope of mild-to-moderate PD vs. HC | P-value: slope of advanced PD vs. HC | P-value: slope of HC |
| --- | --- | --- | --- | --- | --- |
| Sway_PSAC95ER2:logMean | 0.122 | 0.035 | 0.786 | 0.025 | 0.129 |
| Sway_PSAC95ESA:logMean | 0.040 | 0.006 | 0.204 | 0.009 | 0.187 |
| Sway_PSACCMV:logMean | 0.097 | 0.045 | 0.021 | 0.250 | 0.349 |
| Sway_PSACCRMS:logMean | 0.169 | 0.031 | 0.200 | 0.061 | 0.635 |
| Sway_PSACMV:logMean | 0.065 | 0.012 | 0.071 | 0.065 | 0.207 |
| Sway_PSACRG:logMean | 0.124 | 0.022 | 0.679 | 0.032 | 0.092 |
| Sway_PSACRMS:logMean | 0.141 | 0.037 | 0.771 | 0.031 | 0.144 |
| Sway_PSACSMV:logMean | 0.098 | 0.022 | 0.093 | 0.052 | 0.084 |
| Sway_PSACSRG:logMean | 0.103 | 0.028 | 0.991 | 0.032 | 0.067 |
| Sway_PSACSRMS:logMean | 0.112 | 0.031 | 0.673 | 0.019 | 0.126 |
| Sway_PSAN95ER2:logMean | 0.119 | 0.035 | 0.805 | 0.026 | 0.135 |
| Sway_PSAN95ESA:logMean | 0.040 | 0.006 | 0.199 | 0.009 | 0.189 |
| Sway_PSANCRMS:logMean | 0.165 | 0.031 | 0.192 | 0.061 | 0.639 |
| Sway_PSANRMS:logMean | 0.060 | 0.012 | 0.767 | 0.026 | 0.149 |
| Sway_PSANSRMS:logMean | 0.110 | 0.031 | 0.710 | 0.020 | 0.132 |
| TUG_STSD:Mean | 0.223 | 0.050 | 0.867 | 0.331 | 0.538 |
| TUG_TA:logMAD | 0.277 | 0.072 | 0.169 | 0.197 | 0.337 |
| TUG_TD:logMAD | 0.110 | 0.409 | 0.066 | 0.912 | 0.200 |
| TUG_TD:Median | 0.018 | 0.060 | 0.989 | 0.087 | 0.074 |
| TUG_TPV:Median | 0.008 | 0.001 | 0.147 | 0.013 | 0.780 |
| Walk_APAMAA:logMean | 0.179 | 0.031 | 0.549 | 0.267 | 0.947 |
| Walk_GLLC:logMAD_A | 0.052 | 0.600 | 0.474 | 0.018 | 0.568 |
| Walk_GLLC:logMAD_L | 0.007 | 0.708 | 0.524 | 0.012 | 0.922 |
| Walk_GLLC:Median_A | 0.006 | 0.002 | 0.430 | 0.037 | 0.369 |
| Walk_GLLC:Median_L | 0.005 | 0.002 | 0.439 | 0.038 | 0.362 |
| Walk_GLLDS:logMAD_A | 0.015 | 0.009 | 0.095 | 0.015 | 0.081 |
| Walk_GLLDS:logMAD_L | 0.002 | 0.026 | 0.009 | 0.001 | 0.146 |
| Walk_GLLDS:Median_A | < 0.001 | 0.155 | 0.278 | 0.001 | 0.627 |
| Walk_GLLDS:Median_L | < 0.001 | 0.191 | 0.288 | 0.002 | 0.663 |
| Walk_GLLGCD:logSD_A | < 0.001 | 0.083 | 0.641 | < 0.001 | 0.713 |
| Walk_GLLGCD:logSD_L | 0.002 | 0.082 | 0.949 | 0.001 | 0.881 |
| Walk_GLLGCD:Median_A | 0.012 | 0.004 | 0.408 | 0.050 | 0.292 |
| Walk_GLLGCD:Median_L | 0.008 | 0.003 | 0.408 | 0.042 | 0.294 |
| Walk_GLLGS:logSD_A | 0.042 | 0.015 | 0.330 | 0.002 | 0.055 |
| Walk_GLLGS:logSD_L | 0.164 | 0.032 | 0.509 | 0.021 | 0.072 |
| Walk_GLLGS:Median_A | < 0.001 | < 0.001 | 0.559 | < 0.001 | 0.166 |
| Walk_GLLGS:Median_L | < 0.001 | 0.002 | 0.425 | < 0.001 | 0.240 |
| Walk_GLLLSM:Median_L | 0.133 | 0.529 | 0.345 | 0.045 | 0.327 |
| Walk_GLLPIC:logMAD_L | 0.073 | 0.059 | 0.610 | 0.068 | 0.279 |
| Walk_GLLPIC:Median_A | 0.011 | 0.020 | 0.362 | 0.008 | 0.485 |
| Walk_GLLPIC:Median_L | 0.146 | 0.071 | 0.454 | 0.087 | 0.519 |
| Walk_GLLPTO:logMAD_A | 0.009 | 0.048 | 0.137 | 0.001 | 0.837 |
| Walk_GLLPTO:logMAD_L | < 0.001 | 0.002 | 0.083 | < 0.001 | 0.297 |
| Walk_GLLPTO:Median_A | 0.002 | 0.069 | 0.843 | 0.002 | 0.507 |
| Walk_GLLPTO:Median_L | < 0.001 | 0.226 | 0.404 | < 0.001 | 0.212 |
| Walk_GLLS:logMAD_A | 0.132 | 0.490 | 0.618 | 0.025 | 0.887 |
| Walk_GLLS:logMAD_L | 0.002 | 0.014 | 0.626 | 0.006 | 0.260 |
| Walk_GLLS:Median_A | 0.038 | 0.389 | 0.848 | 0.020 | 0.506 |
| Walk_GLLS:Median_L | < 0.001 | 0.288 | 0.026 | < 0.001 | 0.444 |
| Walk_GLLSD:logSD_A | < 0.001 | 0.007 | 0.409 | < 0.001 | 0.915 |
| Walk_GLLSD:logSD_L | 0.002 | 0.088 | 0.486 | 0.001 | 0.769 |
| Walk_GLLSD:Median_A | 0.003 | 0.007 | 0.896 | 0.025 | 0.770 |
| Walk_GLLSD:Median_L | 0.118 | 0.027 | 0.222 | 0.151 | 0.169 |
| Walk_GLLSLE:logSD_A | 0.034 | 0.035 | 0.421 | 0.001 | 0.073 |
| Walk_GLLSLE:logSD_L | 0.040 | 0.004 | 0.441 | 0.022 | 0.078 |
| Walk_GLLSLE:Median_A | < 0.001 | 0.015 | 0.746 | < 0.001 | 0.197 |
| Walk_GLLSLE:Median_L | < 0.001 | 0.029 | 0.493 | < 0.001 | 0.258 |
| Walk_GLLSLS:logMAD_A | 0.275 | 0.075 | 0.362 | 0.222 | 0.390 |
| Walk_GLLSLS:logMAD_L | 0.251 | 0.327 | 0.194 | 0.056 | 0.921 |
| Walk_GLLSLS:Median_A | < 0.001 | 0.239 | 0.047 | 0.001 | 0.551 |
| Walk_GLLSLS:Median_L | 0.034 | 0.298 | 0.786 | 0.017 | 0.461 |
| Walk_GLLSW:logMAD_A | 0.136 | 0.493 | 0.618 | 0.026 | 0.895 |
| Walk_GLLSW:logMAD_L | 0.002 | 0.014 | 0.631 | 0.006 | 0.261 |
| Walk_GLLSW:Median_A | 0.038 | 0.389 | 0.848 | 0.020 | 0.506 |
| Walk_GLLSW:Median_L | < 0.001 | 0.289 | 0.026 | < 0.001 | 0.445 |
| Walk_GLLTDS:logMAD_A | 0.012 | 0.057 | 0.102 | 0.002 | 0.362 |
| Walk_GLLTDS:logMAD_L | < 0.001 | 0.005 | 0.009 | < 0.001 | 0.940 |
| Walk_GLLTDS:Median_A | < 0.001 | 0.208 | 0.179 | < 0.001 | 0.391 |
| Walk_GLLTDS:Median_L | 0.004 | 0.253 | 0.631 | 0.010 | 0.497 |
| Walk_GLLTOA:logMAD_A | 0.018 | 0.714 | 0.304 | 0.012 | 0.974 |
| Walk_GULMV:logMAD_A | 0.069 | 0.250 | 0.970 | 0.059 | 0.410 |
| Walk_GULMV:Median_A | < 0.001 | 0.129 | 0.793 | 0.003 | 0.224 |
| Walk_GULMV:Median_L | 0.027 | 0.312 | 0.770 | 0.015 | 0.432 |
| Walk_GULROM:logMAD_L | 0.086 | 0.147 | 0.389 | 0.458 | 0.742 |
| Walk_TA:Median | 0.181 | 0.265 | 0.055 | 0.194 | 0.274 |
| Walk_TN:Mean | 0.003 | 0.046 | 0.021 | 0.001 | 0.095 |
| Walk_TPV:Median | 0.085 | 0.221 | 0.370 | 0.018 | 0.659 |

### References

Fan, J., and Li, R. 2001. Variable selection via nonconcave penalized likelihood and its oracle properties. *Journal of the American statistical Association* *96* (456):1348-1360.
